# Identification and characterization of differentially co-expressed head-to-head gene pairs in pancreatic cancer

**DOI:** 10.1101/2023.10.14.23297044

**Authors:** Conrad Wang, Chao Ren

## Abstract

Head-to-head (H2H) gene pairs are an evolutionarily conserved genomic configuration (sharing a bidirectional promoter) with implications in cancer. This study investigates the transcriptional mechanisms of H2H genes in pancreatic ductal adenocarcinoma (PDAC). We found that H2H pairs involving housekeeping and tumor suppressor genes maintained a stabler expression correlation, while most pairs had reduced co-expression. Differential co-expression analysis revealed 15 H2H gene pairs significantly altered in PDAC. The gene pairs RPL7-RDH10 and STAC-RNF38 showed both high transcriptional factor similarity and differential co-expression, and enrichment analysis highlighted FOXC1 and YY1 as key regulators of altered H2H pairs. Our characterization of H2H transcriptional patterns in PDAC provides insights into bidirectional promoter disruption. The identified H2H gene signatures offer paired biomarker potential for improved PDAC detection. Further study of mechanisms maintaining H2H stability could spur new therapeutic approaches.

## Introduction

The human genome is thought to be organized in compact, functional modules. One such module is the head-to-head gene pair (H2H genes), an evolutionarily ancient and conserved genomic configuration with implications in human disease. In this arrangement, two adjacent genes are divergently transcribed from a shared bidirectional promoter, leading to higher coordinated expression of neighboring gene pairs through sharing of cis-regulatory sequences (Trinklein et al., 2004). Disruption of this coordinated expression through loss of regulatory elements can have profound effects, particularly for H2H pairs involving crucial genes like tumor suppressors. Cancer has been associated with altered H2H co-expression and disruption of key H2H pairs involved in DNA repair and carcinogenesis. However, mechanisms of differential H2H gene expression in specific cancers remain unclear. This study focuses on characterizing H2H transcriptional patterns in pancreatic ductal adenocarcinoma (PDAC), in order to shed light on promoter disruption mechanisms and identify putative biomarkers. Elucidating conserved H2H regulatory processes disrupted in PDAC can be important for revealing new therapeutic targets and improving early detection gene signatures for this disease. PDAC is one of the deadliest cancers because it is mostly diagnosed in late stage when therapy is extremely difficult and survival rates are slim, and thus it is important to devise new methods for early screening and detection. In this study, we investigate the potential of paired genetic biomarkers for early detection.

H2H gene pairs have been shown to be evolutionarily conserved – particularly in vertebrates – and the genes in each pair implied to have significant cofunction (Li *et al*., 2006). Several H2H genes are known to be co-expressed and function in certain pathways, such as PSENEN and U2AF1L4 in T-cell activity or the collagen genes COL4A1 and COL4A2 (Didych *et al*., 2012). A previous study found that H2H pairs with similar functions have especially high expression correlation (Li *et al*., 2006). Coordinated expression of the two genes has been implied to be biologically significant and efficient, being evolutionarily conserved as an efficient method of gene expression (Li *et al*., 2006).

As the coordinated expression is controlled by a shared bidirectional promoter region, individual regulatory elements can directly influence transcription of both genes. For example, methylation silencing has been found to be effective in co-regulation of these H2H pairs, simultaneously silencing both genes (Shu *et al*., 2006). Transcriptional activation of H2H genes is more frequent than random gene pairs, meaning molecules like transcription factors (TF) have an important role in regulating the promoter regions of the genes (Li *et al*., 2006). Isolated TATA boxes also promote bidirectional transcription *in vitro*, albeit at the expense of the levels of transcriptional output (Bagchi *et al*., 2016). H2H pairs are thus more reliant on regulatory factors, in order to maintain their coordinated expression. Though efficient for their co-expression, this reliance can also be a vulnerability, making them especially susceptible to changes in transcription factor or such regulatory disruptions.

In the context of different cancers, mutations or epigenetic changes to bidirectional promoters can have catastrophic snowball effects on H2H gene transcription, with potential large-scale protein alterations or mutations. Particularly in the context of key genes – e.g., housekeeping genes (HKGs) and tumor suppressor genes (TSGs) – the promotion of tumorigenesis can be devastating. Since H2H genes are enriched with various TFs, they are particularly vulnerable to transcriptional changes. Loss or gain of regulation often leads to differential co-expression of H2H pairs, which can have double the impact than they might have in a single gene promoter. Altered regulatory elements leading to normally co-expressed, individually regulated genes with asynchronous expression and weakened correlation is known as differential co-expression. When gene products form functional complexes, altered expression matrices can affect protein co-production, accelerating cellular processes involved in cancer or other conditions (Choi *et al*., 2005).

As previous pan-cancer analyses of H2H genes have revealed that gene pairs have significantly altered co-expression matrices, it is quite possible that unique biomarkers could be found among differentially co-expressed H2H pairs (Chen *et al*., 2021). H2H genes involving DNA repair and carcinogenesis genes being differentially co-expressed is also rather common, disrupting the expression coordination of these crucial genes. Enhanced in breast cancer, the BRCA1, BRCA2, FANCB2, and FANCD genes involve bidirectional promoters (Yang *et al*., 2007). It’s possible these significantly altered and recognizable H2H genes could serve as putative biomarkers for other cancers, particularly in screenings for early-stage asymptomatic cancers, e.g., pancreatic cancer. This study characterizes differentially co-expressed H2H genes in pancreatic ductal adenocarcinoma (PDAC). We calculate expression, functional, and TF similarity between genes in a pair, and investigate H2H pairs involving key gene types, i.e., tumor suppressor genes. By studying co-expression scores of H2H pairs in healthy and PDAC pancreases, we identify key differentially co-expressed genes, as well as the TFs most directly relevant to them.

## Materials and methods

### Identification of H2H pairs

H2H genes are divergently transcribed on opposite strands, with transcription start site (TSS) coordinates less than 1000 bp apart. The threshold of 1000bp had been adopted by most researchers studying H2H genes, such as Li *et al*. and Trinklein *et al*. The tentative rationale is that genes separated more than 1000bp apart are unlikely to share a bidirectional promoter, since few genes have such long upstream regions. H2H pairs are either overlapping (negative distance between two TSS sites with overlapping promoter regions) and nonoverlapping (with TSS pairs separated with positive distance, no overlapping promoter region). Using these parameters, TSS cluster coordinates for the human genome were downloaded from DBTSS9 (Suzuki *et al*., 2017). After subtracting the reverse strand coordinate from the forward strand coordinate, gene pairs that are on opposite strands and have TSS coordinates < 1 kb apart were determined to be H2H genes. For genes with multiple TSS clusters, the coordinate closest to its neighboring gene was used.

### H2H gene expression in pancreatic cancer

3 public datasets were downloaded from Gene Expression Omnibus/GEO (GSE16515, GSE71989, GSE62165). Each had expression samples from both healthy and cancer patients, allowing for side-by-side comparison between PDAC and healthy pancreas. Expression counts were aligned against the identified H2H pairs.

### Identification of H2H pairs involving special genes

Sequences for several special gene types aligned to H2H genes. Housekeeping genes (HKGs) were downloaded from DBTSS, TSGs were downloaded from TSGene, DNA repair genes downloaded from MD Anderson Cancer Center, epigenetic modifier genes downloaded from Epifactors, Oncogenes downloaded from OncoKB, and angiogenesis genes downloaded from InvivoGen (Suzuki *et al*., 2017; Zhao *et al*., 2015; Wood *et al*., 2001; Marakulina *et al*., 2022; Chakravarty *et al*., 2016; Invivogen).

### Measuring functional similarity

Functional similarity was calculated between the two genes in a pair, by using the GeneSim function in the R package GoSemSim (Yu *et al*., 2010). Presets “combine” as “BMA” and “measure” as “Wang” were used to estimate similarity between two genes. The Wang method weighs GO annotations of genes, prioritizing direct relationships such as “is a” over “part of” (Wang *et al*., 2007). By querying all the GO terms associated with the two genes being compared and weighing them accordingly, it follows the formula

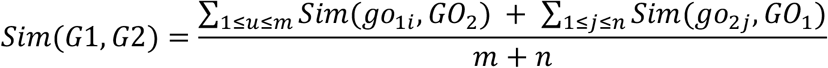

where *go* is any GO term annotated to one of the genes. The Wang method considers the depth of a GO annotation in its ontology hierarchy, whereas older methods like the Resnik method assigns the same weight to all annotations. This approach has been credited as being more specific and accurate, especially for terms further down in ontology hierarchy (Thomas *et al*., 2012).

### Measuring expression correlation

Expression correlation was calculated by finding Pearson Correlation Coefficient (PCC) scores between the expression of each gene in a pair. This was done separately for tumor and healthy samples in each dataset. To account for variance between samples and to enhance PCC accuracy, outliers in each row/gene (defined as counts larger than Q3+1.5IQR (1.5 interquartile ranges larger than quartile 3)) were replaced with the median expression value. Some values in the datasets were ten-fold greater than others, and therefore drastically affected PCC scores and accuracy. Median replacement eliminated these outliers, enhancing the precision of the expression scores.

When comparing the PCC scores of special gene types (like HKGs), random samples were taken of H2H genes of corresponding sample size as comparison. This was done to account for differences in sample size, due to limited H2H pairs with special gene types. P-values were calculated via paired or unpaired t-tests between random and special H2H pairs. Density plot diagrams and violin plots were generated for visual representation of PCC comparison under healthy and cancer conditions.

### Identification of “shifted” H2H pairs

For each dataset, the log_2_ fold change (log_2_FC) of PCC values were calculated for H2H genes under healthy and tumor conditions with the formula 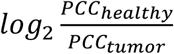. Gene pairs whose log_2_FC values were consistent across all 3 datasets were determined to be differentially co-expressed, consistently having altered co-expression scores (mostly eliminating random fluctuations in expression). Gene pairs with a log_2_FC score above 1.5 were defined as the differentially co-expression pairs, consistently experiencing significant loss- or gain-of-coregulation.

### Calculation of intersected TF

TF enrichment was performed on shifted gene pairs using Enrichr with a manually curated top 10 enriched TFs for each pair; TFs past the first ten generally had high p-values and are thus insignificant. The relevance/significance of each enriched TF was evaluated based on its p-value and whether it was enriched to both genes at the same time. TFs enriched to both genes indicates strong evidence of binding to their bidirectional promoter, as TFs binding there likely coordinate the co-expression of both genes. A formula was created for calculating the total intersected TF and relevance score for TF binding evidence in a bidirectional promoter

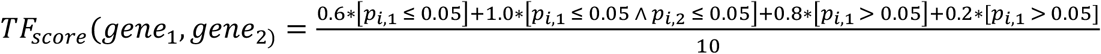

where *Pi*.1 refers to the p value of TFs relevant to one gene, and *Pi*.1 ^*Pi*.; refers to the p values of a TF with binding evidence to both genes. The highest weighted score (1.0) was assigned to TFs significantly enriched in both genes in a pair, indicating strong regulation evidence of the shared bidirectional promoter. The lowest score (0.2) indicates that all 10 TFs were not enriched to only 1 gene in the pair.

### Statistical analyses

All statistical analyses were done using Python. Packages *numpy, pandas* were used for normalization or calculating PCC. Packages *seaborn* and *matplotlib* were used to generate the figures, including violin diagrams and density plots. Unless otherwise specified, all p-values were calculated via Mann-Whitney U test. For example, in calculating the p-value for functional similarity, we opted for an unpaired t-test to compare any random H2H pair with any random gene pair, since the data was overall quite normally distributed.

## Results

### Identification of human H2H gene pairs

1646 protein coding H2H genes were identified, selecting genes which are on opposite strands to avoid counting adjacent gene pairs. This updated list of protein coding H2H gene pairs is a 38% increase from the most recent pan-cancer analysis of H2H pairs, which identified 1196 protein coding pairs (Chen *et al*., 2021). Updates to genome annotation databases and TSS data can account for this increase.

Since H2H configurations can have both positive and negative TSS distances – negative TSS distances indicating overlap between TSS regions (Fig 1A) – we compared the TSS distance distributions of the 1646 pairs: results are shown in Fig 1B. Considering transcriptional direction of 5’ to 3’, distances were calculated by subtracting the coordinates of the reverse strand from the forward strand (Fig 1A).

**Figure 1.**
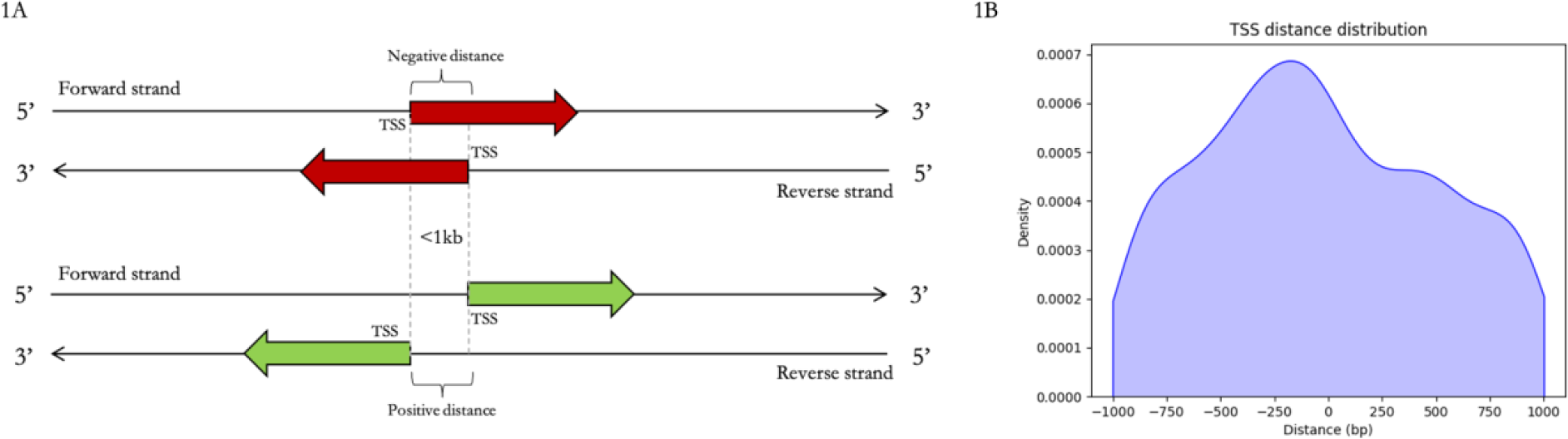
H2H TSS distances. (A) Schematic diagram of H2H organization. (B) TSS distance distribution in H2H pairs

The distributions of H2H pair distances were different from previous research from Li, *et al*., (2006) and Chen, *et al*., (2021) (Li *et al*., 2006; Chen *et al*., 2021). Chen et al., (2021) found that a large portion of protein coding H2H genes were separated by distances of 1-400 bp, whereas our data suggests a prominent range of -400 to 0, indicating most of our H2H pairs have overlapping promoter regions. Still, these results can be explained by the mechanisms of shared bidirectional promoters. Overlap of promoter regions in a bidirectional promoter can be spatially efficient, creating a densely packed H2H unit. In either case, the majority of H2H pairs are spaced ±250-400 base pairs apart. This might provide insights into the length of bidirectional promoters, suggesting that the region is more often around 250-400 bp long. The Eukaryotic Promoter Database assumes that human promoters are generally no longer than 500bp, since its promoter data is truncated at 500bp upstream of the transcription start site. As such, our results indicating a main clustering of H2H pairs spaced -250-400 bp long appears to be reasonable, and may yield insights into the clustering and length of bidirectional promoters.

### Functional similarity between H2H genes

Our functional semantic similarity of H2H genes are consistent with earlier studies, showing that similarity of function is not significantly favored in H2H pairs. Function GeneSim from the GOSemSim R package was used with the Wang method, scaling the similarity of two genes between 0 and 1. Of the 1194 H2H pairs that had GO terms matched to both genes, the mean GeneSim score was 0.54, and the median 0.495.

When comparing the distribution of functional similarity scores of H2H pairs with randomly paired genes, a t-test was conducted and no significant difference was found (p= 0.60). Though not strictly normally distributed, the data overall has an apex around 0.5, and its slight normal trend was considered when picking t-test to validate statistical difference. Since the sample size was sufficiently large (1194), the statistical results are accurate. Fig 2 models the distribution of similarity scores. These results are corroborated by Chen, *et al*., (2021). Even though some H2H pairs like COL4A1-COL4A2 form complexes, it appears that there is no evolutionary selective pressure to necessarily group co-functional genes together. Potentially, this could be a mechanism to minimize damage caused by mutations: theoretically, damage to a bidirectional promoter can silence both genes, leading to a complete lack of a protein complex that might be able to function partially with only one unit.

**Figure 2.**
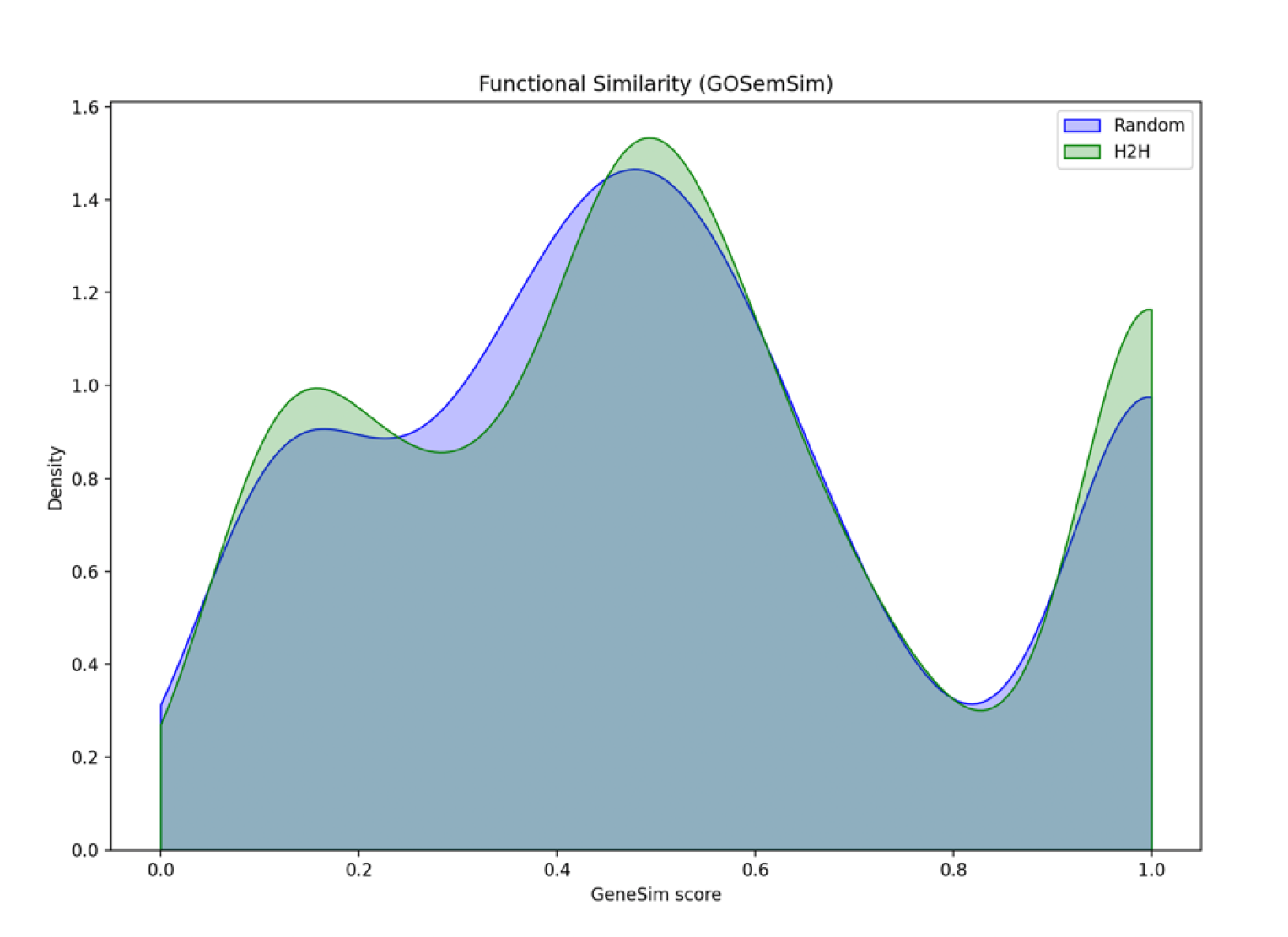
Distribution of GOSemSim functional similarity scores in H2H and random gene pairs, with equal sample sizes for each group. No significant difference between two groups (p>0.10), with both groups having peaks around 0.1, 0.5, and 1.0

Something to consider is how GoSemSim defines and calculates functional similarity. Split into cellular functions, molecular functions, and biological process, the relationship between genes are weighed and considered. Strong relationships like “is-a” are weighed with a higher arbitrary score like 1.0, while weaker relationships like “part-of” are weighed lower like 0.4. By tracing the function of each gene with a series of “is-a” and “part-of” relationships to a larger category of biological processes and comparing the two, GoSemSim can calculate the similarity between two gene terms. It’s important to clarify that high functional similarity scores like 0.8 or 1.0 don’t always indicate direct relationships like in a co-functional unit (using HIST1H2BG-HIST1H2AE as an example again), but can have many diverse roles that contribute to a common function.

The functional similarity comparison suggests that throughout the evolution and conservation of H2H pairs, there are limited factors favoring functionally similar genes to be grouped together in a pair. Nonetheless, functional modules such as BRCA1-NBR2, which act somewhat complementarily (relevance to cell cycle regulation), are arranged H2H, but their functions are not exactly similar.

### Expression similarity of H2H pairs in healthy and cancer conditions

Pearson Correlation Coefficient scores (PCC) were used as indices for co-expression scores between the two genes in a pair, and we found that H2H pairs in tumor had significantly lower PCC scores. Absolute value of the PCC scores were taken before calculating mean and median, to model the overall strength of expression similarity. As expected, the median PCC values of healthy gene pairs were much higher, with mean 0.335 and median 0.310. Because of altered regulatory molecules leading to differential co-expression of the pairs, tumor sets had only a mean of 0.187 and a median of 0.156. Violin plots for the distribution of these values are shown in Fig 3, for each of the datasets.

**Figure 3.**
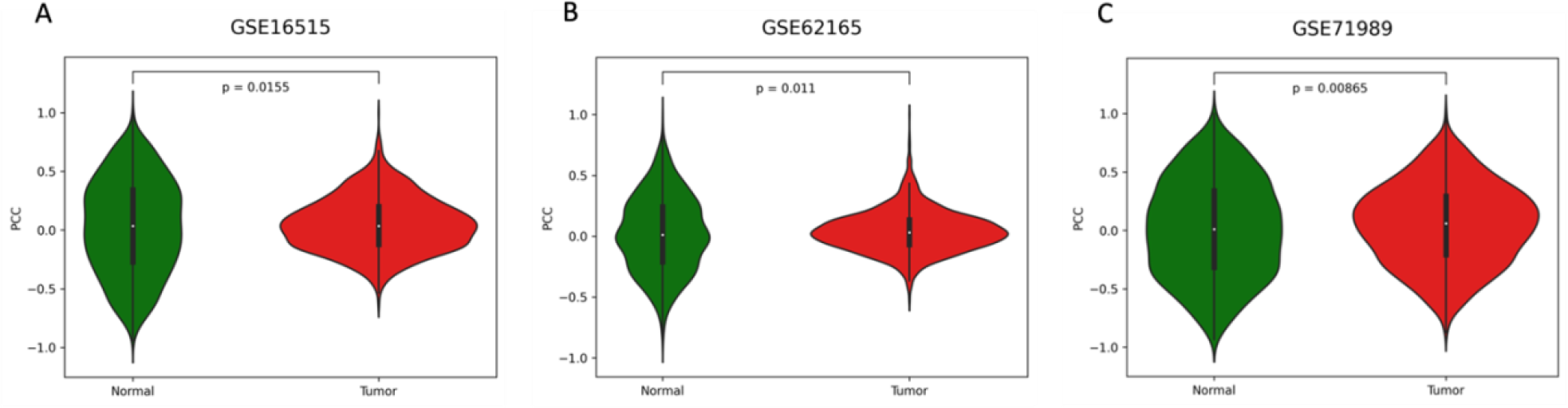
Violin plot comparing H2H co-expression distribution under healthy and PDAC conditions, where healthy pancreas shown in green and tumor pancreas in red. Measured using PCC scores. P values between two groups were calculated using paired t-test. Each dataset had +1000 samples, indicating a healthy sample size and thus reliable t-test results.

### Expression correlation of H2H pairs involving special gene types

Apart from studying general H2H genes and random gene pairs, we also investigated the co-expression of H2H genes involving gene types with specific roles in the cell, such as HKGs or TSGs. Housekeeping genes are stably and consistently expressed in all cell types, involved in core cellular processes, such as metabolism, DNA replication, transcription, and translation (Eisenberg & Levanon, 2013). TSGs, DNA repair genes, and oncogenes have prominent roles in cancer progression; mutations or failures in such genes cascade into catastrophic genomic abnormalities, resulting in tumors and cancer. Other gene types – angiogenesis genes and epigenetic modifiers, for example – are similarly involved in cancer processes, albeit to a lower degree. After aligning these special gene types in H2H pairs, we identified special pairs with 1 or 2 of these key gene types, shown in Table 1.

**Table 1:** H2H pairs involving special gene types, through aligning key gene lists with H2H lists. H2H pairs where both genes were of a gene type were also identified. HKG had the largest degree of involvement in H2H pairs.

|  | Total | Occurs > once | Occurs twice | % arranged H2H |
| --- | --- | --- | --- | --- |
| Housekeeping (HKG) | 522 | 160 | 6 | 30.7% |
| Tumor suppressor (TSG) | 1217 | 149 | 4 | 12.2% |
| DNA repair | 219 | 42 | 0 | 19.2% |
| Angiogenesis | 141 | 29 | 0 | 20.6% |
| Epigenetic modifier | 801 | 139 | 3 | 17.4% |
| Oncogene | 332 | 56 | 1 | 16.9% |

We found that 160 out of 522 (31%) HKGs downloaded from DBTSS were part of an H2H gene, with 6 of them being HKG-HKG pairs. Considering how approximately 60-70% of HKGs are protein coding, 31% is reasonable, since Chen, *et al*., (2021) found that 35% of all H2H pairs involved HKGs. There is a less consistent pattern of TSGs and DNA repair genes being arranged head-to-head. 149 out of 1217 TSGs (12%) and 42 out of 219 DNA repair genes (19%) were arranged head-to-head, which is insufficient to prove any significant advantage of these genes being favored in H2H pairs. There is little straightforward rationale or advantage of arranging TSGs and DNA repair genes in H2H configurations (Eisenberg & Levanon 2013; Chen *et al*., 2006).

As HKGs are known to be stably expressed across all cell types, we examined the expression matrices of H2H pairs involving them. Fig 4 shows the distribution of PCC scores under healthy and tumor conditions for H2H genes with and without HKGs. Under healthy conditions, there is no significant difference between PCC scores of HKG and non-HKG (p=0.32). Though unclear from the graph, there seems to be a significant difference between PCC distributions of H2H genes with HKGs under tumor conditions. Values are spread out more, with a Gaussian curve slightly skewed to the right. Statistical comparison through a t-test revealed the p-value to be 7.14e-06. The low p-value indicates that the difference is highly unlikely to be due to chance, indicating a significant difference between the two groups; with higher co-expression matrices in tumors compared to non-HKG pairs. Despite the sample size for HKGs being small compared to the entire set of H2H pairs, we calculated paired t-test multiple times in the cancer condition of HKG-H2H pairs, and every trial had a p value below 0.05 and around 0.01, showing significant difference.

**Figure 4.**
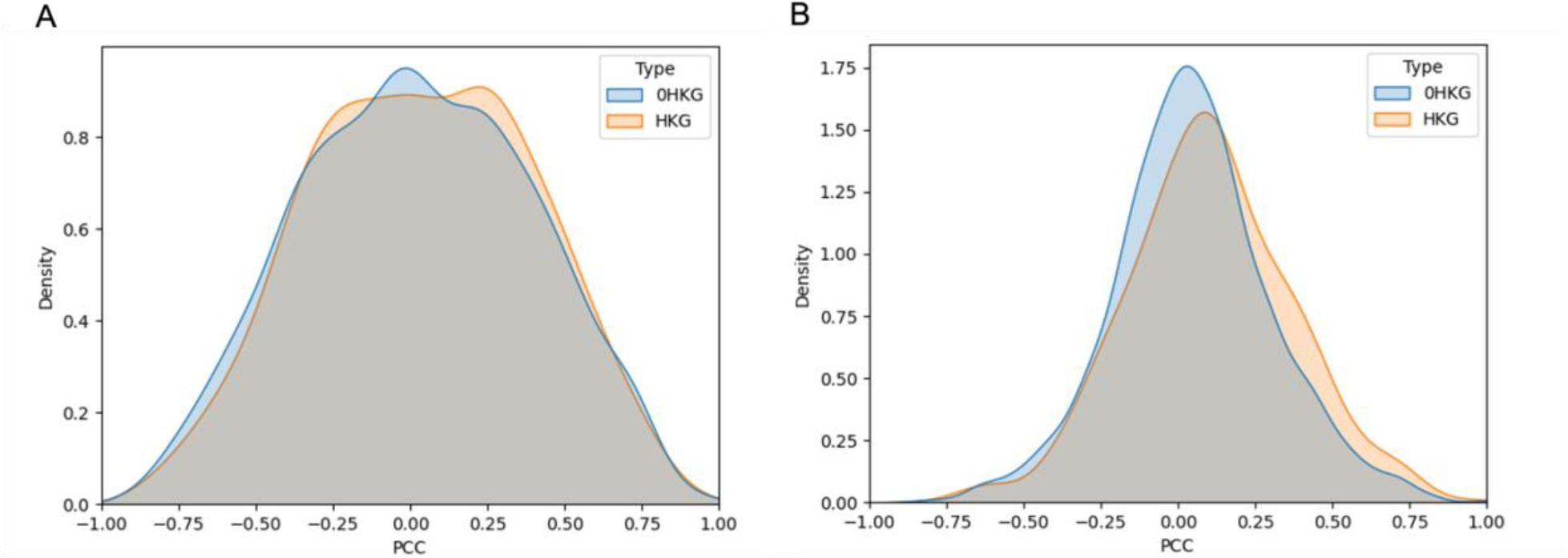
comparison between PCC scores of HKG gene pairs. (A) Comparison between H2H pairs involving 0 or at least 1 HKG in healthy pancreas expression data. (B) Comparison between H2H pairs involving 0 or at least 1 HKG in tumor pancreas expression data

Knowing that co-expression scores of HKG genes could be significantly more stable compared to non-HKG genes, we also investigated the PCC scores of other main gene types, such as tumor suppressor genes.

Unlike HKGs, the expression matrix of TSG-involved H2H pairs are significantly different from other H2H pairs even under normal conditions. As shown in Figure 5A, the median PCC score of HKG-H2H pairs has a more negative value than non-TSG H2H pairs (p<0.0005). The data indicates the H2H pairs involving TSGs have stronger expression matrices. Past research has identified key TSG-H2H gene pairs with significant cofunction. For example, Xu, *et al*., (1997) identified tumor suppressor gene BRCA1 as being an H2H gene with non-coding gene NBR2, which induces hepatoblastoma cell malignancy and cell proliferation (Xu *et al*., 1997; NCBI). Especially when forming functional complexes with other genes (like the histone pair HIST1H2BG-HIST1H2AE), higher co-expression seems to be encouraged in TSG-H2H pairs.

**Figure 5.**
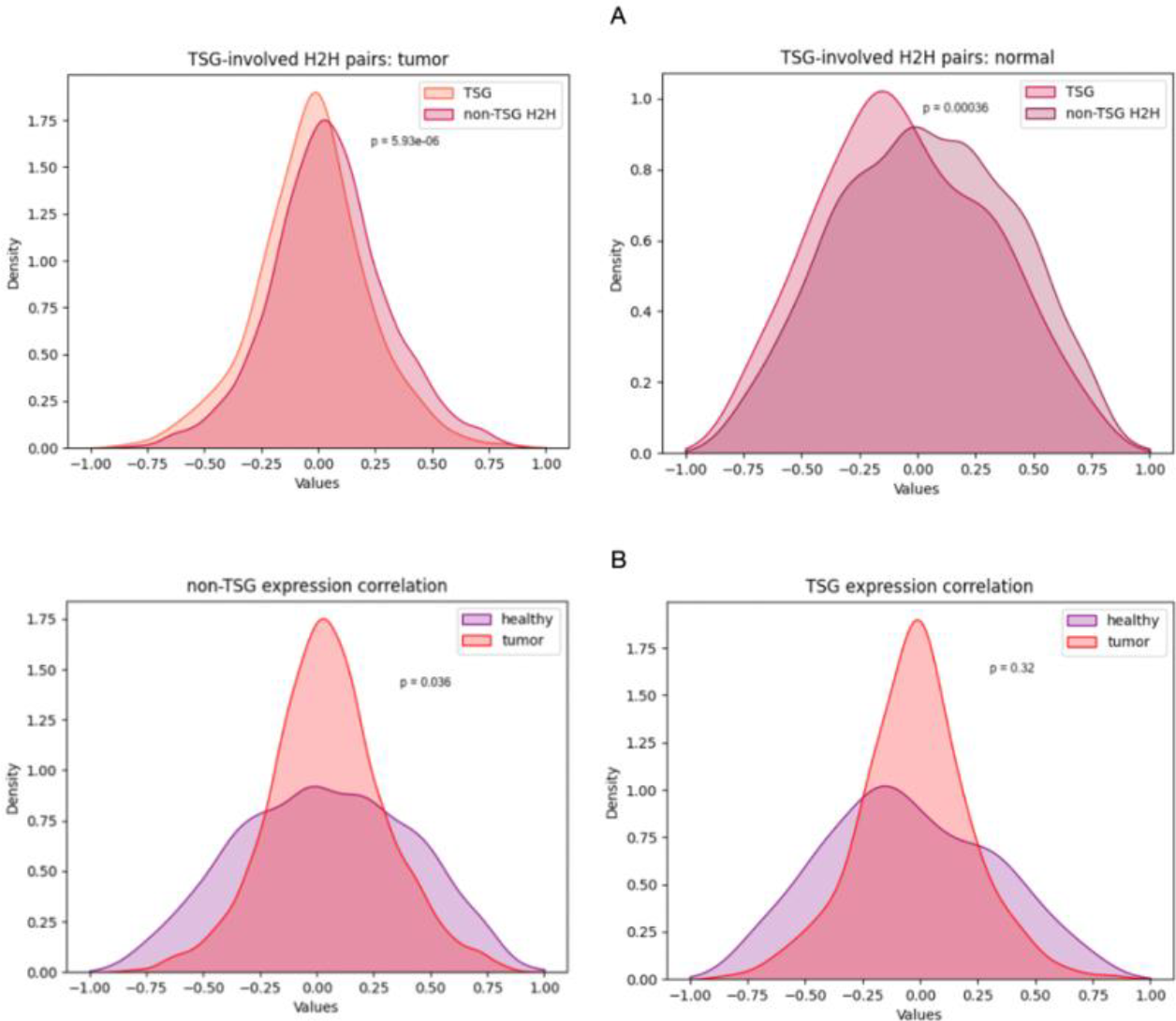
PCC scores of H2H pairs with and without tumor suppressor genes. (A) Distribution of PCC scores of H2H pairs without TSGs under healthy and tumor conditions. (B) Distribution of PCC scores of H2H pairs containing at least 1 TSG under healthy and tumor conditions

Under tumor conditions, the distribution of both TSG and non-TSG H2H pairs are both Gaussian curves, showing a rather even distribution of values. However, the distribution of the TSG-involved H2H pairs is skewed slightly to the left, with mean and median PCC values being slightly negative (thus a stronger correlation between the two pairs). We found the upper and lower bound of the middle 40% of the data: {-0.14, 0.07} for TSG-involved, and {-0.08, 0.16} for non-TSG. Judging from the shape of the distribution curves, TSG-involved H2H pairs seem to be slightly more resistant to differential co-expression and altered expression coordination.

To further explore the relative stability of TSGs, we compared expression correlation of TSG and non-TSG H2H genes under healthy and tumor conditions, shown in Fig 5B. For both TSG and non-TSG pairs, co-expression scores are much lower in tumor conditions, mostly clustering -0.25 to 0.25. However, a t-test result revealed that healthy and tumor PCC scores were not significantly different in the TSG-involved H2H pairs, despite the graph seeming so. With a p-value of 0.32, co-expression matrices for TSG-involved H2H pairs remain statistically constant under healthy and tumor conditions.

However, we encourage verification of our results concerning TSGs. Because the sample size of TSG-involving H2H pairs was rather small compared to other gene types (149 pairs with at least 1 TSG), p-values calculated via unpaired t-test might deviate between trials. Due to this inaccuracy, we ran the code calculating unpaired t-test multiple times, with the results ranging from p=0.3 to p=0.5, still suggesting little significant difference between the PCC logFC values of TSG-involved H2H pairs and random H2H pairs.

Driven by the results from HKGs and TSGs, we compared the PCC scores of DNA repair genes under healthy and tumor conditions. Consistent with the data from HKGs and TSGs, there is also no significant difference in PCC scores of tumor and healthy H2H pairs when they involve DNA repair genes (p = 0.13, not significant). Data is shown in Figure 6. However, a possible concern with this data is the very limited number of H2H pairs involving DNA repair genes. Given a small sample size, a p-value of 0.13 might be caused by simply not having enough data. Observing Figure 6B, we noted that more DNA repair H2H genes in cancer seemed to have a PCC score clustered closer to 0, possibly due to silencing of DNA repair genes during the mutational and growth stage of cancer.

**Figure 6:**
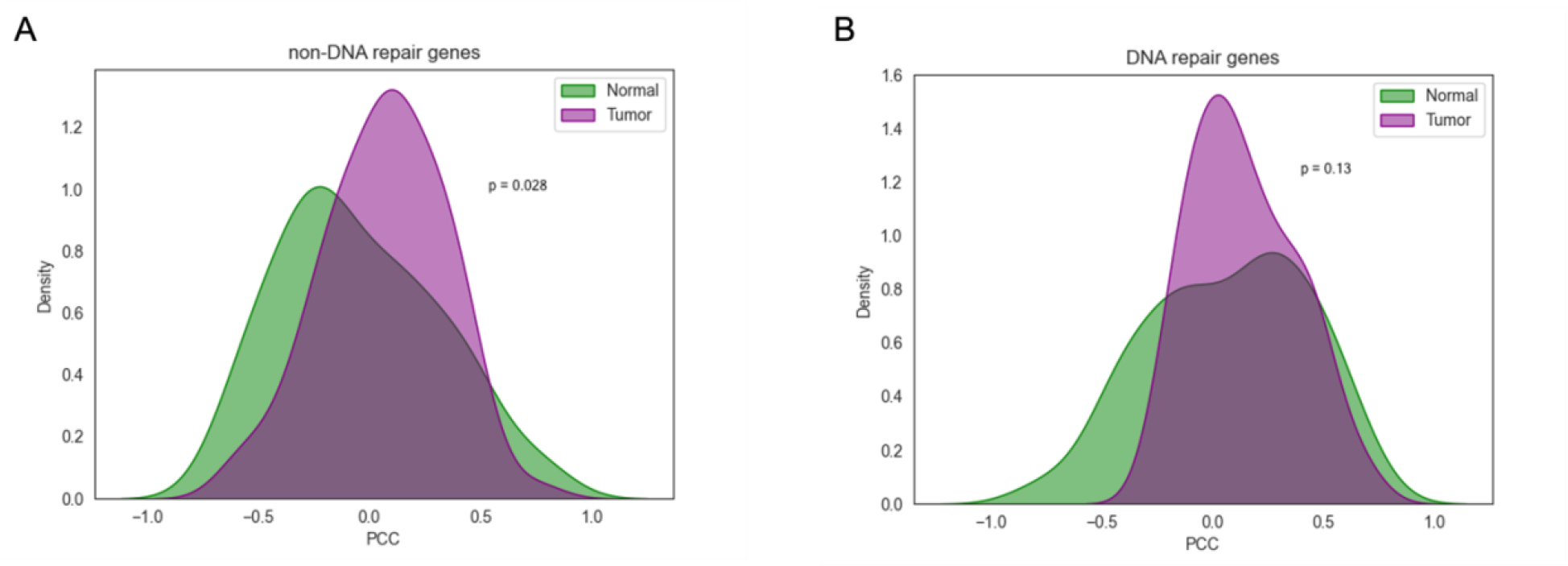
PCC score distribution under healthy and tumor conditions of H2H pairs (A) without DNA repair genes (B) containing at least 1 DNA repair gene

In profiling the expression similarity of H2H genes involving angiogenesis genes, epigenetic modifier genes, and oncogenes, there was no reason to expect any significant stability compared to random H2H genes (unlike HKGs). Because of the smaller sample size of these genes, even t-tests on random H2H pairs of the same sample size yielded insignificant results.

Taking the mean fold change values of PCC scores in healthy and tumor conditions and plotting them in density diagrams, we found that angiogenesis genes (Figure 7A), epigenetic modifier genes (Figure 7B), and oncogenes (Figure 7C) did not demonstrate the exceptional co-expression stability demonstrated by HKGs. Whereas the mean fold change values of random H2H pairs were 0.903, angiogenesis genes had a mean of 0.877, epigenetic modifier genes 1.01, and oncogenes 1.159. Again, it is important to address the sample sizes for these special gene types. For angiogenesis, epigenetic modifier, and oncogenes, there are only 48, and 99

**Figure 7.**
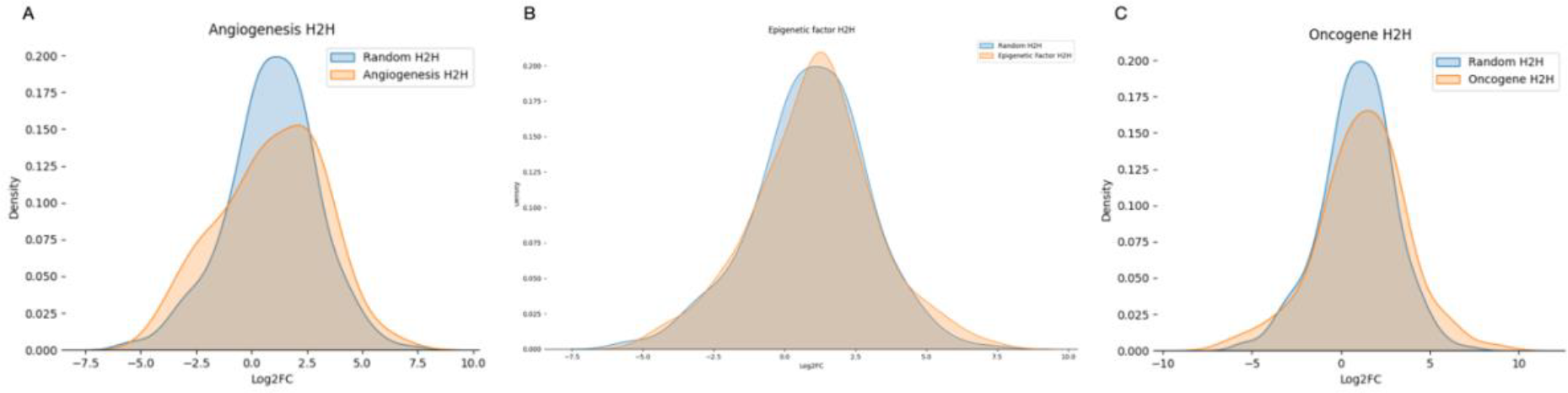
Log2 fold change of PCC scores in random H2H pairs versus H2H pairs involving (A) angiogenesis genes (B) Epigenetic modifier genes (C)Oncogenes

**Figure 8.**
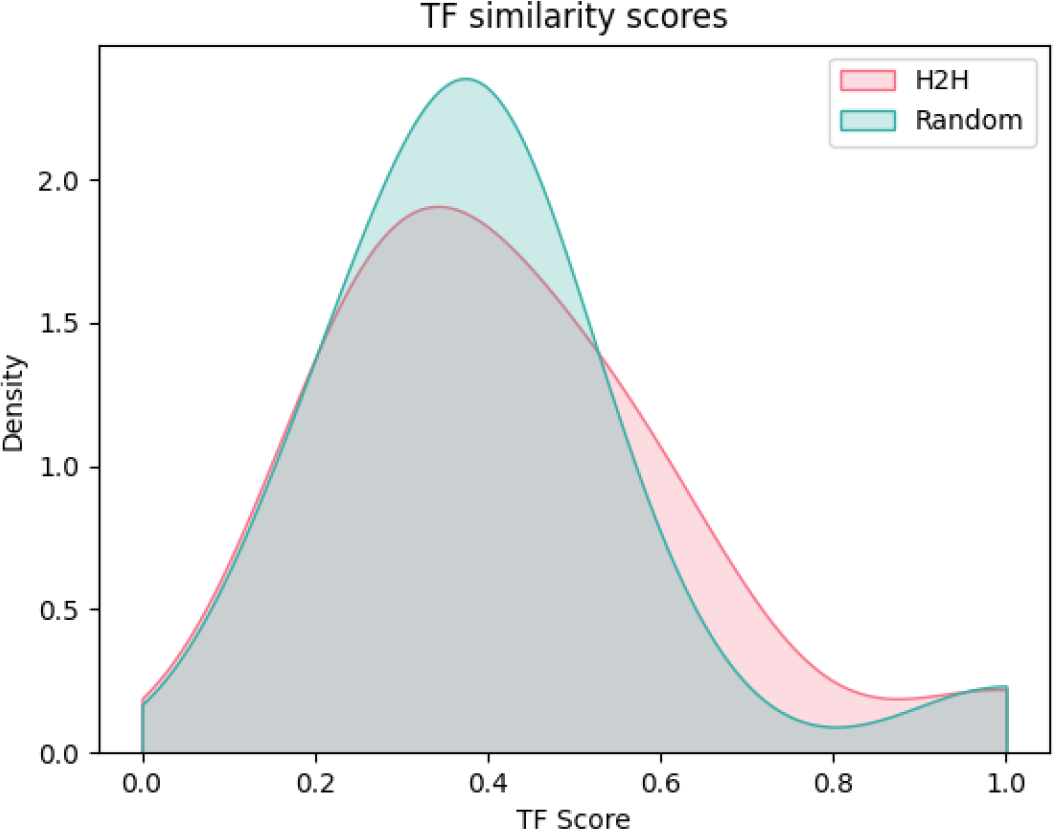
TF similarity scores of H2H and random pairs, calculated with weights prioritizing TFs significantly enriched in both genes in an H2H pair

### Identification of differentially co-expressed (DCG) H2H genes

From our statistical analysis, we identified 15 key differentially co-expressed H2H pairs in PDAC. Differentially co-expressed gene pairs (DCGs) are H2H genes whose co-expression matrix are significantly altered; under healthy conditions, the bidirectional promoter coordinates the co-expression of both genes, using shared TFs to maintain transcriptional stability. This is a key feature of H2H genes, allowing for the stable and efficient dual transcription matrix. But with random transcriptional changes associated with pancreatic cancer, many of these co-expression complexes are disrupted, with each gene being transcribed independently.

To identify DCG pairs, we calculated the log fold change (logFC) of PCC scores under healthy and tumor conditions with 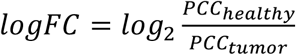 . The assumption was that if a gene pair had consistent logFC values across the three datasets (where logFC score was either consistently positive or consistently negative), then PDAC had a non-negligible influence on their co-expression matrix. 241 of the 1646 pairs were consistently altered, with 15 of them having logFC values above 1.5 or below -1.5 for all samples (thus a 2.8 fold change in PCC score). The arbitrary logFC threshold of 1.5 was chosen to identify the most significantly altered H2H pairs. Since only 15 pairs had logFC values greater than 1.5, the threshold was defined to select only the most altered pairs. Since the goal of the study is to sift through the H2H pairs to find the pairs with strongest potential as biomarkers, defining the arbitrary logFC threshold as 1.5 is one way to identify the strongest candidates.

Some of the DCG pairs with highest logFC scores include DERL3-SLC2A11, ETFA-FAM47E, and LPAR-TMA7. The logFC scores of these pairs are clustered around 4-5, indicating a 16- to 32-fold change in their co-expression scores.

### Transcription factor similarity of DCG H2H pairs

After enriching the 15 significant DCG pairs with Enrichr and then calculating their significant TF similarity scores, the three H2H pairs with the highest TF similarity scores were RPL7-RDH10, STAC-RNF38, and DERL3-SLC2A11. They had the TF similarity scores 1.0, 0.64, 0.60, respectively. The 1.0 score for RPL7-RDH10 means that of the 10 top annotated TFs for the pair, all 10 of them had significant binding evidence to both genes, indicating an extremely high level of TF similarity. As these three genes are not only differentially co-expressed but also have large degrees of TF similarity, we concluded that these three pairs are especially involved in H2H bidirectional formation, and are affected by PDAC.

To evaluate the effect of shared TFs on H2H pair co-expression, we also calculated the PCC scores of these three gene pairs of interest and compared them to average PCC scores. We found that the geometric mean of the absolute value PCC scores was 0.181, a relatively weak correlation overall. In comparison, the geometric means of the three gene pairs was much higher, being 0.571, 0.413, and 0.419 respectively. RPL7-RDH10, having the highest number of shared TFs, naturally had the highest expression correlation.

Of all TFs with binding evidence to the H2H pairs, FOXC1 and YY1 were frequently enriched for both genes in differentially co-expressed pairs, suggesting they may be associated with disruption of coordinated expression. FOXC1 is also overexpressed in tumor conditions, with a 47% expression increase.

## Discussion

In this study, we examined differentially co-expressed H2H pairs in pancreatic cancer, including the expression profiles of different types of gene pairs. From the expression similarity data, we found that co-expression correlation between general H2H pairs is significantly decreased by pancreatic cancer, with p-values around 0.01 for all three datasets (p=0.0155, 0.011, 0.00865, as shown in Fig 3). We also found that among the key gene types, HKGs and TSGs were the most stably expressed, the differences between PCC scores in healthy and tumor samples being insignificant. Using the change in PCC scores between healthy and tumor samples for each gene pair, we then identified 15 key H2H gene pairs with logFC of PCC greater than 1.5, with 11 of them experiencing significant loss-of-co-regulation. Using these insights coupled with TF annotation data from Enrichr, RPL7-RDH10 was identified as the most significant differentially co-expressed H2H pair, with exceptional TF regulation similarity between the two genes. Of those shared TFs, FOXC1 and YY1 appear multiple times among the differentially co-expressed pairs.

Overall, the pervasive differential co-expression of H2H pairs in PDAC underscores how bidirectional promoters efficiently couple key genes but are vulnerable in disease. The data-driven identification of TFs like FOXC1 as frequent enriched regulators of disrupted pairs provides direct evidence of this mechanism. By linking concrete H2H pair examples like RPL7-RDH10 to general trends, these results elucidate principles of bidirectional promoters in cancer while nominating actionable biomarkers.

Though this study only examined the expression of H2H gene pairs in pancreatic cancer, the significance of YY1 and FOXC1 as TF regulators of differentially co-expressed pairs may be generalized to other cancers. Literature has indicated that both TFs are heavily suspected to interact with cancer, since overexpressin of YY1 is correlated with poor cancer clinical outcomes, and “deregulation of FOX proteins is also involved in the development and progression of tumors”. Both TFs are known to, under healthy circumstances, regulate crucial processes like cell proliferation, transcriptional control, and apoptosis (Khachigian *et al*., 2018; Han *et al*., 2017). Knowing that these two TFs are correlated with many cancers, and misregulate differentially co-expressed H2H pairs in pancreatic cancer, our findings might spur future research into the relationship between YY1 or FOXC1 and head to head gene pairs in a broader context.

As hypothesized, the expression correlation between H2H pairs is much lower in cancerous samples than in healthy samples. These results were expected, given that regulation and coordination of the bidirectional promoters are heavily reliant on TFs and other fragile regulatory mechanisms. Because bidirectional promoters are often enriched with hypersensitive DNase and modified histones, H2H genes are often in an active state, prone to co-expression disruptions and modifications (Li *et al*., 2006). When regulation mechanisms are disrupted in PDAC, the delicate matrix is disrupted and two genes are left to their own independent promoters (reflected in a much lower PCC score). This poses the greatest risk to functionally complementary pairs, or H2H genes that form complexes together and are heavily reliant on being co-transcribed and their proteins produced in similar amounts. For example, the histone pair HIST1-H2BG and HIST1-H2AE has their expression matrix consistently disrupted, potentially causing issues with DNA compacting.

Of all the key gene types, HKGs seemed to be the most involved in H2H gene configurations. These results show that HKGs are more resistant to regulatory disruptions, being stably and consistently co-expressed in their bidirectional promoters. HKGs have been exerted extra selective pressure to be organized in H2H gene configurations, with bidirectional transcription maintaining the fundamental functions of HKGs and some complementary genes (Lin et al., 2007; Chen et al., 2021). The results enforce this conclusion, demonstrating the relative stability of H2H pairs when involving HKGs, possibly a reflection of evolutionary pressures to maintain the security of crucial housekeeping genes.

The relative stability of HKGs by themselves is not surprising, given their consistent expression in all cell types, among vastly different genetic environments in different disorders or cancers. Being foundational, HKGs have long faced evolutionary pressures to remain stable and consistently regulated, so their robustness in PDAC is not unexpected. However, data demonstrates the co-expression stability of the HKG-involved H2H genes. Most the pairs involved only 1 HKG, implying that the presence of an HKG could also promote the stability of its H2H gene complement. It may act as an anchor, or through certain mechanisms prioritize the coordination of bidirectional pairs where they reside. Further research into the specific molecular and transcriptional mechanisms of HKG H2H genes will be needed to fully understand these anchoring mechanisms, as well as comprehensive characterization of these neighboring genes that benefit from HKG stability.

Among the other genes with certain implications in cancer, like TSGs or angiogenesis genes, we found that TSGs and DNA repair genes were slightly more resistant to differential co-expression. Synthesizing our previous data about HKG-involved pairs, H2H genes seem to be significantly stable for crucial genes like HKGs TSGs, indicating their role in promoting genetic stability. These results may provide insights into prioritization mechanisms of H2H genes, maintaining stability of key genes to ensure proper coordination.

However, other genes – angiogenesis, epigenetic modifier genes, and oncogenes – demonstrated no significant difference from random H2H genes. The mean logFC values of their expression scores in healthy and tumor groups deviated roughly around 0.9 for the first two and 1.15 for oncogenes, which aren’t significantly different from the logFC values of random H2H genes. It wasn’t surprising that most of the logFC values across these three types of genes were vaguely clustered around 0.8 to 1.2, indicating that co-expression in healthy conditions tended to be 1.7-2.3 times that in tumor. As shown from the violin plots, PDAC had a non-negligible effect on various of these co-expression mechanisms, which tended to separate the transcription of genes in a pair and cause differential co-expression. After all, there are no significant features of these gene types (unlike the evolutionarily encouraged stability of HKGs) for them to be resilient to transcriptional modifications.

From the threshold of differentially co-expressed H2H gene pairs having consistent logFC scores above 1.5 or below -1.5, most differentially co-expressed gene pairs experience losses of coordination rather than gain of coordination. Mutations and transcriptional changes are much more likely to disrupt delicate bidirectional promoter matrices, severing correlation links between two genes in a pair. Of the 15 significant DCGs we identified, only 4 of them had negative values, corresponding to a strengthening of expression correlation. The overwhelming majority of weakened expression correlations suggests that loss of regulation tends to prevent H2H gene co-expression, leading to disorder.

In the TF similarity analysis on significant DCG pairs, we found that the range of scores to be 0.2-1.0. This suggests that H2H genes have slightly higher TF similarity compared to random gene pairs, with a higher percentage of them having multiple TFs with binding evidence to both genes; this is due to the shared bidirectional promoter region in H2H genes.

TF enrichment analysis on these DCG pairs identified FOXC1 and YY1 as commonly enriched in shifted H2H genes. FOXC1 has been associated with tumor development and metastasis, reenforced by its role in DCG pairs (Han *et al*., 2017). Examining expression data of FOXC1, we also found it was consistently over-expressed in pancreatic cancer, meaning higher concentrations of FOXC1 can be detrimental to gene expression coordination. Though YY1 was not indexed in any of our three datasets, much literature has cited YY1 as being instrumental in epithelial-mesenchymal transition (EMT) and tumor progression in various cancers (Palmer *et al*., 2009; Han *et al*., 2017).

In the last step of our TF enrichment analysis, all 30 DCG pairs were fed into Enrichr. We found that SMAD4 was one of the top enriched TFs. Coincidentally, SMAD4 was also the third mutated gene in PDAC differential expression analysis, bested only by the classic cancer genes KRAS and TP53.

Lin, *et al*., (2007) identified 7 key TFs overrepresented in bidirectional promoters: NF-Y, Nrf-1, YY1, GABP, MYC, E2F1, and E2F4 (Li *et al*., 2006). Of these 7 overrepresented TFs, 5 were enriched to the differentially co-expressed pairs in PDAC. YY1, one of the key TFs identified in this analyses, was enriched in three out of the 15 pairs; MYC and E2F1 were enriched in two of the pairs; E2F4 and NRF1 were both enriched once. These TFs may act as determinants of transcriptional direction, not just passively associated with bidirectional promoters (Bagchi *et al*., 2016). Presence of these bidirectional-inducing TFs in differentially co-expressed pairs show how fragile TFs can be; though capable of inducing bidirectional and coordinated expression, they are also particularly vulnerable to over- and under-regulation.

Overall, TF similarity analysis yielded three key insights. TF similarity of H2H pairs is indeed higher than that in random pairs, with a higher percentage of the pairs sharing numerous significantly enriched TFs. RPL7-RDH10 was identified as the top DCG H2H pair, having exceptional TF similarity and also being drastically affected by PDAC, with mean log2FC of 2.12. Lastly, FOXC1 and YY1 were identified as key TFs in DCG H2H genes.

Inevitably, there could be external factors that affect our findings in co-expression and transcriptional factors similarity. For the top H2H pairs like RPL7-RDH10, we also investigated whether there could be functional relationship between the two that would be advantageous for coordinated expression, like some cellular pathway requiring the protein products of both genes. However, none of the top differentially co-expressed pairs had any notable similarity or shared processes, at most dealing with the same type of biomolecular (like proteins).

There are some assumptions that had to be made when dealing with H2H pairs that must be addressed in this paper. The most important assumption involves the definition and identification of differentially co-expressed gene pairs. Knowing that H2H gene pairs are co-expressed bidirectionally, there are two main ways that their co-expression could be altered. In the first case, both genes in the pair are significantly up- or down-regulated, with regulatory elements shifting the expression of both genes the same way (simultaneous over-expression or silencing of the gene pairs). In the second case, the co-expression matrix between the two genes are changed, with their expression being shifted differently. For example, one gene could be up-regulated while the other is down-regulated. Our study focuses mainly on differential co-expression (the second case), and not altered co-expression. This decision was made since several H2H pairs, like HIST1H2BG-HIST1H2AE, need both genes to be stably expressed so that the resulting protein complex can work properly. In differential co-expression, HIST1H2BG might be under-expressed while HIST1H2AE is over-expressed, resulting in limited numbers of functional histone complexes. As such, the approach we opted for was investigating the Pearson Correlation Coefficient between the two genes: lower PCC scores indicated loss of co-expression and thus differential co-expression.

The accuracy of genome studies relies heavily on the quality and consistency of input data. In our study, we only selected datasets GSE16515, GSE62165, and GSE71989 since those were the few publicly available datasets that included expression profiles from both healthy and PDAC patients. Many GEO datasets had PDAC expression available, but lacked a comparison with healthy patients. The selection of these datasets can account for the variance between different datasets, like demographics, apparatus, data processing etc. In our calculations, change of expression scores were only compared *within* such a dataset (we would not compare healthy expression scores of GSE16515 with cancerous expression scores of GSE62165).

Another consideration is the variance between patients in the same dataset. Due to external factors like inherited mutations unrelated to PDAC, or simply inaccurate expression readings, there are some values in the expression scores that are significantly different from the other values. Of the expression scores of a single gene in a dataset, there could sometimes be values tenfold greater than the other values. Including such outliers could have a large impact on the final PCC value, with extreme outliers decreasing PCC significantly. We accounted for these discrepancies by eliminating the outliers. Since these outliers were rare and most likely random, the assumption was that they were completely unrelated to H2H gene co-expression or PDAC. Thus all outliers 1.5 inter-quartile-ranges above the third quartile (see Materials & Methods) were pruned from the datasets to ensure most realistic PCC scores.

In addition, the sources of the three datasets GSE16515, GSE62165, and GSE71989 are from Mayo Clinic, Vlaams Instituut voor Biotechnologie, and University of Florida, respectively. Given that the location of these sources differ, it is important to consider possible biases between the datasets. Despite all three datasets originating from Caucasian-dominated countries, it should still be kept in mind that the patient demographic could differ between the datasets and samples.

This study could be improved with a large sample size. A plan to conduct meta-analysis on PDAC expression data is in progress, where larger amounts of expression profiles of both tumor and healthy pancreas samples are compared. Furthermore, the best way to confirm the role of key TFs in these differentially co-expressed gene pairs is through ChIP-seq analysis on healthy and PDAC pancreas tissue. Only this way can we definitively conclude the change in TF regulation of a specific H2H gene; as of now we rely on inferring by examining TF binding data to healthy H2H genes, then calculating the change in expression levels of that TF. A better alternative would be to inspect TF-gene binding in PDAC, identifying key TFs in not only DCG pairs, but possibly other gain-of-regulation of loss-of-regulation H2H genes.

## Author contributions

Conrad Wang designed, carried out the study, and wrote & edited the paper. Chao Ren wrote the code. All authors have read and approved the final manuscript.

## Abbreviations

H2H –: head-to-head
PDAC: pancreatic ductal adenocarcinoma
DCG –: differentially co-expressed gene
HKG: housekeeping genes
TF: transcription factor
PCC: Pearson correlation coefficient

## Acknowledgement

Special thanks to Yuan-Yuan Li, from Shanghai Center for Bioinformation Technology, and Yunqin Chen, from the School of Life Sciences and Biotechnology from Shanghai Jiao Tong University for guidance on head-to-head genetics, and for suggesting the topic. Special thanks to Lo Sosinski from Michigan State University for providing valuable guidance with R and bioinformatics, and for editing/revising the final paper. The authors thank them for their advice.

## Data availability statement

Original data and figures presented in the study are all included in the paper. The datasets analyzed during the current study are available in the Gene Expression Omnibus, from datasets GSE16515, GSE62165, and GSE71989. Further inquiries for individual files and code can be directed to the corresponding authors, and original files will be included in supplementary materials.

